# Early life and Adulthood Depression: Birth seasonality on demographic characteristics of depressive symptoms

**DOI:** 10.1101/2020.07.14.20153361

**Authors:** Hao Zhou, Danni Peng-Li, Juan Chen, Dong Sun, Bin Wan

## Abstract

**Background:** Environmental and biological factors in early-life in-utero can have critical health implications in adulthood. However, despite the extensive literature on the association between depressive symptoms and female gender, higher age, lower education, and lower socio-economic status, no studies have thus far investigated these depression-related demographic factors in connection with early-life environment. Here, the present study aimed to explore the effects of birth seasonality on demographic characteristics of depressive symptoms in adults.

**Methods:** We employed data from the project of Chinese Labour-forces Dynamic Survey (CLDS) 2016, containing the epidemiological data of depressive symptoms with a probability proportional to size cluster and random cluster sampling method in 29 provinces of China with final sample of 16,181 participants was analysed. Logistic regression analyses were performed to test the relations between having depressive symptoms and various demographic characteristics in the overall population and different layers driven by the season of birth (Spring: March, April, and May; Summer: June, July, and August; Autumn: September, October, and November; Winter: December, January, and February).

**Results:** In line with previous studies, female gender, higher age, lower education, lower satisfaction of family income, and northern geographical region were the depressive symptoms-related factors. Season of birth did not significantly contribute to having depressive symptoms. Gender and satisfaction of family income were significant for the linear trend in people born in spring, summer, autumn, and winter. Age was significant for linear trend in people born in spring and winter. Education was only significant in spring. The categorical variable of geographical region had different associations with depressive symptoms across seasons of birth.

**Conclusions:** Our findings indicate that although season of birth is not significantly associated with depressive symptoms, it influences the effects of the demographic factors on depressive symptoms (particularly in age). The present study sheds lights on the discussion of environmental and biological effects related to season of birth on adult mental health.

## 1. Introduction

Depression refers to a group of mental disorders characterized by significant mood depression. Approximately 264 million people worldwide are affected by depression (1, 2) and have experienced a significant decrease in quality of daily life (3, 4). Besides, robust evidence have shown that depression is statistically associated with various kinds of diseases (5-7). Since the Reform and Opening in China, the living standards of residents have been greatly improved. However, a series of problems have followed during the urbanization process (e.g. narrow living space, fast-paced life, high-intensity work and heavy life pressure). Consequently, a growing number of people face the disturbance of anxiety and depressive disorders (1, 8), which in turn has caused economic loss and social burden in China (9, 10).

A great amount of research has linked depressive symptoms to socio-demographic factors. For example, being female (vs male) and older (vs younger) as well as having lower (vs higher) income level and education degree is followed by a higher risk of experiencing depression (11-14). Likewise, geographic factors also be connected to depression, with individuals in Northern China being more susceptible (15). Even, early environment in-utero can potentially form a long-term impact on human development (16-18). In particular, numerous studies have reported effects of birth seasonality on depressive symptoms. An early report demonstrated that major depression had an excess in spring births in the US (19). In an Italian population, individuals born during spring or summer months were reported with higher sensitivity to seasonal changes (20). In addition, a polish study indicated that birth month might be significantly associated with the course of recurrent depressive disorders (21). Specifically, these scholars suggested that the season of birth was an important factor in adulthood depression.

However, even if there is a clear relation between adulthood depressive symptoms and season of birth, the link may not be direct. For instance, one study found that there were excess births from winter to early spring with mood disorders, particularly in females (22), suggesting that the relation of birth month on depressive symptoms is only prevailing in specific groups. In the same vein, and interaction of birth month and life-style was found to be significant in committing suicide in a Greenlandic population (23). Another researcher found that Australian individuals born in the Southern hemisphere from September to November had the higher prevalence of depressive symptoms compared to individuals born in the Northern (24). Collectively, these studies might shed light on the possible mechanisms of birth effects on depressive symptoms. Birth seasonality may interfere the demographic factors on adulthood depressive symptoms.

Nevertheless, the research thus far is generally confined to data from developed countries and with varying results from the different populations. Recent evidence highlighted the association between season of birth and schizophrenia in a large-scale Chinese study (25), yet evidence on the association between adulthood depressive symptoms and birth seasonality is still lacking. To testify such birth seasonality effects, we explored the influence of season of birth on depressive symptoms in Chinese adults. Further, the individuals were categorized into four groups based on the season of birth (i.e., spring, summer, autumn, and winter). We focused on the relation between demographic characteristics on depressive symptoms across populations with the different birth seasonality, including gender, age, economic status, education degree, and geographic regions.

## 2. Methods

### 2.1. CLDS sample

The data used in this paper were obtained from the China Labour-force Dynamics Survey (CLDS) held by the Centre for Social Science at Sun Yat-sen University in Guangzhou, China (please refer to http://css.sysu.edu.cn for more information about the CLDS data). The sampling method and details were introduced in our former report (26). Shortly, multiple stage-systematic and probability proportionate to size (PPS) sampling methods were employed. The surveyors responsible were trained strictly to fulfil the requirements of the project.

### 2.2. Measurements

#### Demographic characteristics

The demographic categorical variables in our analyses included gender, age, education degree, socio-economic status, and geographical regions.

- **Gender** was categorized into male and female.
- **Age** was categorized into four groups, i.e., young adults group (from 18 to 30 years), young-middle adults group (from 31 to 45 years), middle adults group (from 46 to 60 years), and elderly group (more than 60 years).
- **Education** as “the highest level of education obtained” was differentiated into three levels including elementary education, secondary education, and higher education.
- **Socio-economic status** was replaced by satisfaction of family income level (low, middle-low, middle, middle-high, high) because this investigation was conducted according to the unit of community and family. In terms of individual’s report, satisfaction of family income was more representative especially regarding mental health. Hoebel and colleagues (27) found that lower objective socio-economic status and lower subjective socio-economic status were independently associated with current depressive symptoms, and that there was a significant indirect relationship between objective socio-economic status and depressive symptoms through subjective socio-economic status.
- **Geography** was clustered into seven regions in China according to the administrative division, including Central, North, East, South, Northwest, Northeast, and Southwest.

#### Depressive symptoms

The depressive symptoms were self-reported by using the Chinese edition (28) of the Centre for Epidemiologic Studies Depression (CES-D) Scale (29). The CES-D is a well-validated screening tool for depression and has been used in many population-based epidemiologic studies worldwide (29, 30). CES-D contains 20 items about symptoms describing the frequency in a past week on a 4-point Likert scale (ranging from 0 to 3) and the higher marks indicate the severer symptoms. The Chinese version of the CES-D scale has showed acceptable reliability and validity among all age groups both in urban and rural populations. Greater than 15 scores were the recognized value of having depressive symptoms (15, 26, 31, 32).

#### Season of birth

Season of birth was categorized based on the common perception of Chinese. Spring is from March to May, summer from June to August, autumn from September to November and winter from December to February. To make the results more comparable, we also selected the representative month for the corresponding season of birth, i.e., the middle month of each season. In the secondary analyses, April represented spring, July represented summer, October represented autumn, and January represented winter.

### 2.3. Statistical modelling

In the whole sample, 21,091 individuals participated in the survey. There were no records of birth month in 3,949 individuals and age in 19 individuals. We also excluded 472 subjects with age below 18 years old. Besides, there were no record of the CES-D scale in 449 persons and education degree in 17 persons. After the screening procedure, the final valid sample size consisted of 16,185 individuals, whose data were included in the statistical analyses.

Firstly, univariate chi-square tests were applied to examine the difference of demographic characteristics and season of birth in contribution to having depressive symptoms. Secondly, we used chi-square tests to examine the difference of demographic characteristics in distribution of having depressive symptoms in individuals with different season of birth. Moreover, we carried out logistic interaction models to test the interaction between demographic variables and birth seasonality on depressive symptoms.

To further investigate independent associations, four multivariate binary logistic regression models were carried out corresponding to each season of birth (i.e. one for spring, summer, autumn, and winter) in order to predict depressive symptoms. Each logistic regression model included gender, age, satisfaction family income, education, and geographic regions as the independent variables. The dummy variable with the lowest proportion of having depressive symptoms in the univariate tests was set as the reference. The estimates used in the model were the odds ratio (OR) and 95% confidence interval (CI). In addition, as for the rank variables including gender, age, satisfaction of family income and education, we presented the *p* for trend to show its linear significance in the logistic models. All statistical tests were performed on IBM Statistic Product and Service Solutions (SPSS) software 22.0 and the significance level was set at 0.05 with two-tailed test. Secondary analyses were performed to test the sensitivity of the present results with regard to season of birth. We selected the middle month of the season of birth as the targeted populations.

## 3. Results

### 3.1 Primary outcomes

As listed in **Table 1**, 7548 (46.6%) males completed the survey. The age of individuals ranged from 18 to 83 with the mean age of 46.3 (SD = 13.3) years. 17.7% individuals were categorized as having depressive symptoms. Female and higher age were significantly associated with having depressive symptoms (all *p*s < 0.001). The satisfaction of family income (χ^2^ = 597.29, *p* < 0.001) and the highest education degree (χ^2^ = 71.96, *p* < 0.001) were associated with having depressive symptoms. In addition, geographic region was significantly related to depressive symptoms (χ^2^ = 48.07, *p* < 0.001), of which the rate of having depressive symptoms peaked in Northeast China and bottomed in South China. Therefore, in the logistic regression models, male, 18-30 years, high satisfaction of family income, higher education degree, and South China, which showed a low rate of having depressive symptoms, would be set as the corresponding category of references.

**Table 1.** Demographic characteristics

| Variables | Non-depressive symptoms |  | Depressive symptoms |  | Chi-square | *p* |
| --- | --- | --- | --- | --- | --- | --- |
|  | N | Proportion | N | Proportion |  |  |
| **Gender** |  |  |  |  | **51.54** | **< 0.001** |
| Male | 6392 | 84.6% | 1160 | 15.4% |  |  |
| Female | 6935 | 80.3% | 1698 | 19.7% |  |  |
| **Age (years)** |  |  |  |  | **44.80** | **< 0.001** |
| 18 - 30 | 2239 | 85.5% | 381 | 14.5% |  |  |
| 31 - 45 | 3649 | 83.9% | 698 | 16.1% |  |  |
| 46 - 60 | 5376 | 81.1% | 1253 | 18.9% |  |  |
| > 60 | 2063 | 79.7% | 526 | 20.3% |  |  |
| **Highest education degree** |  |  |  |  | **71.96** | **< 0.001** |
| Elementary | 9190 | 80.7% | 2199 | 19.3% |  |  |
| Secondary | 2367 | 86.2% | 379 | 13.8% |  |  |
| Higher | 1770 | 86.3% | 280 | 13.7% |  |  |
| **Satisfaction of family income** |  |  |  |  | **597.29** | **< 0.001** |
| Low | 592 | 60.0% | 395 | 40.0% |  |  |
| Middle-low | 2272 | 75.0% | 754 | 25.0% |  |  |
| Middle | 4652 | 83.8% | 900 | 16.2% |  |  |
| Middle-high | 4256 | 87.2% | 622 | 12.8% |  |  |
| High | 1556 | 89.3% | 186 | 10.7% |  |  |
| **Geographic region** |  |  |  |  | **48.01** | **< 0.001** |
| South China | 2791 | 85.0% | 492 | 15.0% |  |  |
| North China | 1282 | 82.3% | 275 | 17.7% |  |  |
| Northeast of China | 871 | 79.5% | 224 | 20.5% |  |  |
| East China | 3346 | 84.0% | 635 | 16.0% |  |  |
| Central China | 1687 | 81.0% | 396 | 19.0% |  |  |
| Southwest of China | 1311 | 80.2% | 324 | 19.8% |  |  |
| Northwest of China | 2039 | 79.9% | 512 | 20.1% |  |  |
| **Season of birth** |  |  |  |  | **4.72** | **0.194** |
| Spring | 3265 | 82.0% | 718 | 18.0% |  |  |
| Summer | 3354 | 81.5% | 760 | 18.5% |  |  |
| Autumn | 3536 | 83.2% | 713 | 16.8% |  |  |
| Winter | 3172 | 82.6% | 667 | 17.4% |  |  |

Regarding seasonality, we observed no significant association with depressive symptoms (χ^2^ = 4.72, *p* = 0.194). 24.6% of the individuals were born in spring (summer: 25.4%, autumn: 26.3%, and winter: 23.7%, **Table 1**).

**Fig. 1** illustrates the proportion of having depressive symptoms in individuals with different demographic characteristics across the four season of birth groups. The trends for having depressive symptoms in gender and satisfaction of family income were stable across the four seasons of birth (**Fig. 1A and D**). However, age and education were not stable in season of birth in summer (**Fig. 1B and C**), and geographic regions had a different trend for having depressive symptoms among the four seasons of birth (**Fig. 1E**).

**Fig 1.**
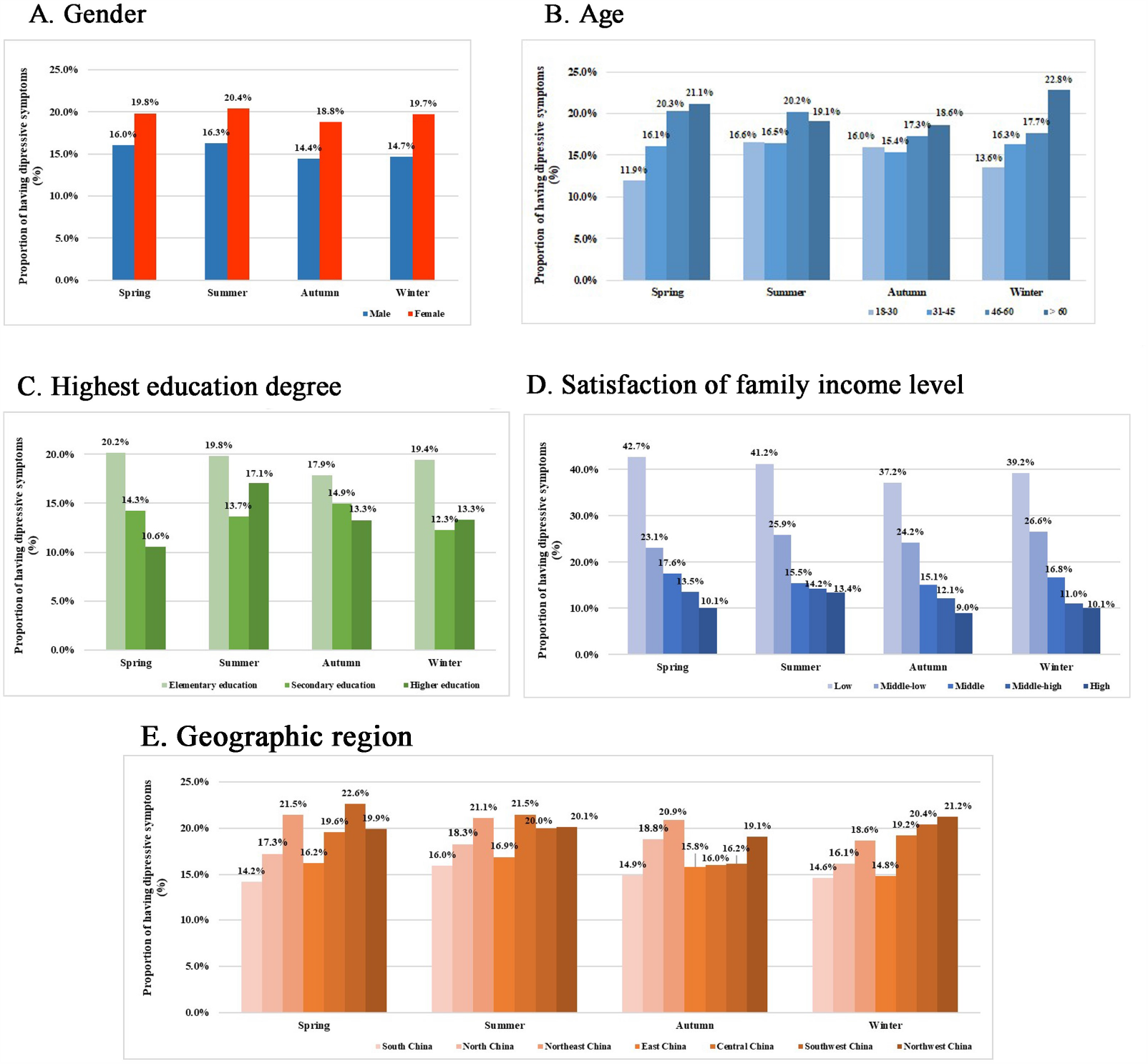
The proportion of having depressive symptoms in individuals with different demographic characteristics across four seasons of birth. (A) Gender and proportion of depressive symptoms; (B) Age and proportion of depressive symptoms; (C) Highest education degree and proportion of depressive symptoms; (D) Satisfaction of family income level and proportion of depressive symptoms; (E) Geographic region and proportion of depressive symptoms.

### 3.2 Interaction of demographic characteristics with season of birth

To test whether there were interactions between season of birth and demographic characteristics on having depressive symptoms, we performed generalized linear analyses with binary response (i.e., depressive symptoms “yes/no” = one of the demographic characteristics + season of birth + interaction). There was no significant interaction of season of birth with gender or satisfaction of family income. The significant interaction effects (age, education, and geographic region) are depicted in **Fig. 2** (including the interaction OR, which set the reference of 18-30 years for age, higher education for highest education degree, South China for geographic region, as well as the reference of each season of birth).

**Fig 2.**
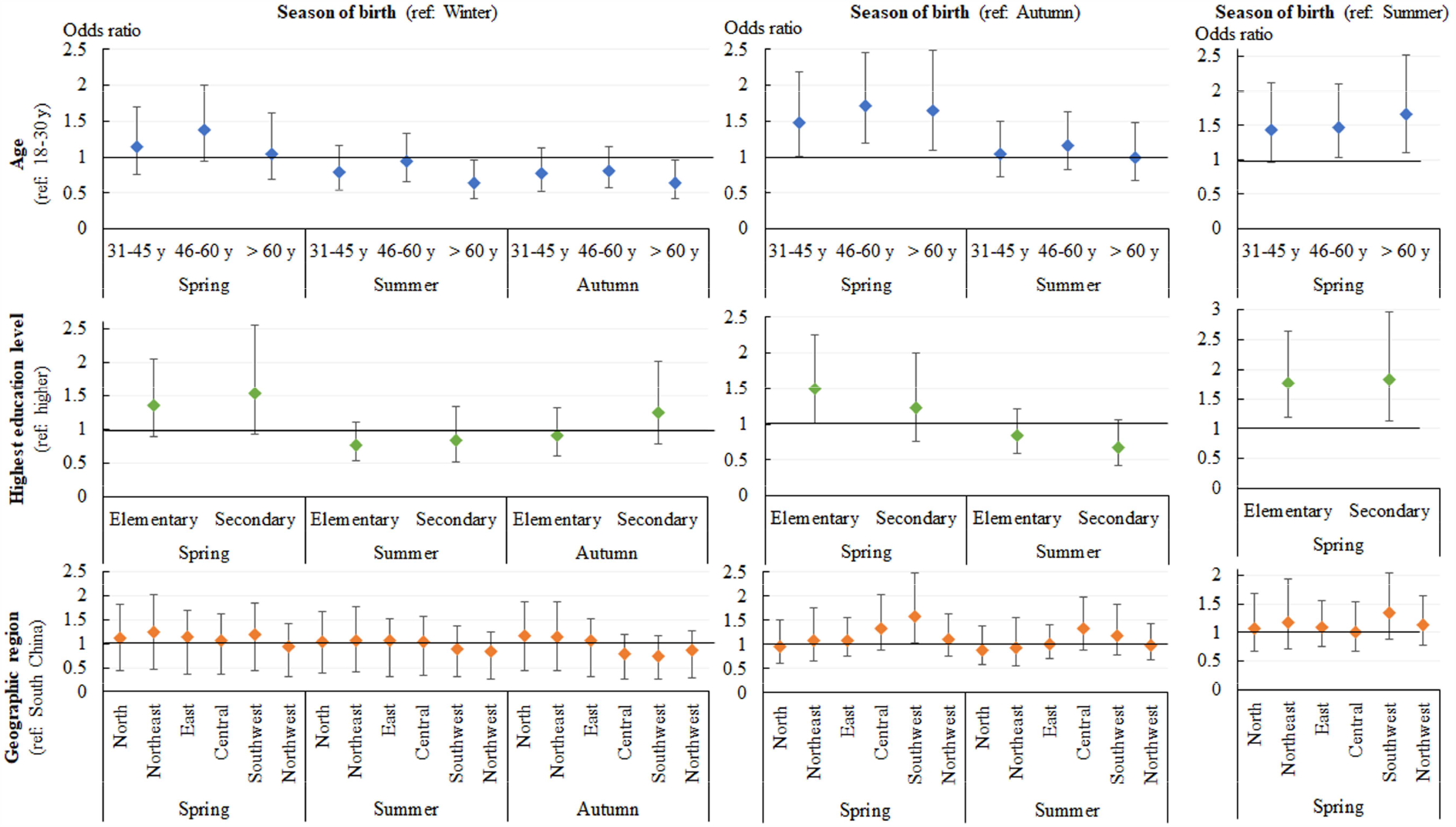
The interaction of season of birth with age, highest education degree, and geographic region on having depressive symptoms. The reference for age, highest education degree, and geographic region was 18-30 years, higher education, and South China respectively. The comparison for season of birth contained 6 types. Ref: reference. y: years.

Specifically, within age susceptibility to depressive symptoms, compared with born in winter, born in summer (OR = 0.63, 95%CI = 0.42, 0.95, *p* = 0.029) and autumn (OR = 0.64, 95%CI = 0.42, 0.96, *p* = 0.032) decreased the risk in individuals aged > 60 years. Born in spring (vs autumn) enhanced the likelihood in individuals aged 31-45 years (OR = 1.49, 95%CI = 1.01, 2.19, *p* = 0.046), 46-60 years (OR = 1.71, 95%CI = 1.19, 2.45, *p* = 0.004), and > 60 years (OR = 1.65, 95%CI = 1.09, 2.49, *p* = 0.018). Born in spring (vs summer) increased the risk in people aged 46-60 years (OR = 1.47, 95%CI = 1.03, 2.10, *p* = 0.033) and > 60 years (OR = 1.66, 95%CI = 1.10, 2.51, *p* = 0.016).

Within education susceptibility, born in spring increased the likelihood to have depressive symptoms, in persons with elementary education compared to born in autumn (OR = 1.50, 95%CI = 1.00, 2.26, *p* = 0.049) and summer (OR = 1.78, 95%CI = 1.20, 2.64, *p* = 0.004), as well as in persons with secondary education compared to born in summer (OR = 1.83, 95%CI = 1.13, 2.96, *p* = 0.014). Within geographic region, born in spring (vs autumn) increased the risk of having depressive symptoms in population living in southwest China (OR = 1.60, 95%CI = 1.02, 2.49, *p* = 0.039).

### 3.3 Multivariate binary logistic regression

**Table 2** shows the multivariate association between demographic characteristics and depressive symptoms across the four layers of season of birth. As expected, the linear associations (*p* for trend) in gender and satisfaction of family income were significant with *p* values lower than 0.001. Compared with males, females were more likely to have depressive symptoms regardless of seasonality. Lower (vs higher) satisfaction of family income indicated a higher proportion of having depressive symptoms (details in **Table 2**).

**Table 2.** Multivariate logistic regression to predict having depressive symptoms across four seasons of birth.

| Variables | Spring | Summer | Autumn | Winter |
| --- | --- | --- | --- | --- |
|  | OR adjusted (95%CI) | OR adjusted (95%CI) | OR adjusted (95%CI) | OR adjusted (95%CI) |
| **Gender** |  |  |  |  |
| (ref: male) | *p* for trend < 0.001 | *p* for trend < 0.001 | *p* for trend < 0.001 | *p* for trend < 0.001 |
| Female | **1.35 (1.14, 1.60)\*\*\*** | **1.35 (1.14, 1.59)\*\*\*** | **1.39 (1.17, 1.64)\*\*\*** | **1.47 (1.23, 1.75)\*\*\*** |
| **Age** |  |  |  |  |
| (ref: 18-30 y) | *p* for trend < 0.001 | *p* for trend = 0.027 | *p* for trend = 0.089 | *p* for trend < 0.001 |
| 31-45 y | 1.08 (0.80, 1.47) | 0.81 (0.61, 1.07) | 0.80 (0.61, 1.05) | 1.00 (0.74, 1.34) |
| 46-60 y | **1.48 (1.11, 1.97)\*\*** | 1.08 (0.83, 1.40) | 0.93 (0.71, 1.22) | 1.12 (0.84, 1.48) |
| > 60 y | **1.70 (1.23, 2.36)\*\*** | 1.10 (0.80, 1.51) | 1.10 (0.81, 1.50) | **1.77 (1.27, 2.45)\*\*** |
| **Education** |  |  |  |  |
| (ref: Higher) | *p* for trend = 0.022 | *p* for trend = 0.027 | *p* for trend = 0.567 | *p* for trend = 0.017 |
| Elementary | **1.47 (1.05, 2.05)\*** | 0.96 (0.73, 1.27) | 1.17 (0.88, 1.56) | 1.12 (0.82, 1.51) |
| Secondary | 1.13 (0.77, 1.65) | **0.69 (0.50, 0.96)\*** | 1.05 (0.75, 1.47) | 0.76 (0.53, 1.09) |
| **Satisfaction of family income** |  |  |  |  |
| (ref: High) | *p* for trend < 0.001 | *p* for trend < 0.001 | *p* for trend < 0.001 | *p* for trend < 0.001 |
| Low | **7.46 (4.90, 11.36)\*\*\*** | **4.98 (3.39, 7.28)\*\*\*** | **6.65 (4.37, 10.11)\*\*\*** | **6.25 (4.10, 9.52)\*\*\*** |
| Middle-low | **2.74 (1.89, 3.98)\*\*\*** | **2.38 (1.72, 3.28)\*\*\*** | **3.51 (2.42, 5.08)\*\*\*** | **3.31 (2.31, 4.75)\*\*\*** |
| Middle | **2.00 (1.40, 2.84)\*\*\*** | 1.25 (0.92, 1.71) | **1.94 (1.35, 2.79)\*\*\*** | **1.90 (1.35, 2.69)\*\*\*** |
| Middle-high | 1.38 (0.96, 1.98) | 1.08 (0.79, 1.49) | 1.43 (0.98, 2.07) | 1.10 (0.77, 1.58) |
| **Geographic region** |  |  |  |  |
| (ref: South China) |  |  |  |  |
| North China | 1.33 (0.95, 1.87) | 1.21 (0.88, 1.67) | 1.35 (0.98, 1.87) | 1.18 (0.82, 1.70) |
| Northeast China | **1.84 (1.29, 2.62)\*\*\*** | 1.44 (0.98, 2.11) | **1.64 (1.14, 2.35)\*\*** | 1.32 (0.91, 1.90) |
| East China | **1.35 (1.02, 1.78)\*** | 1.27 (0.99, 1.64) | 1.28 (0.99, 1.65) | 1.22 (0.92, 1.62) |
| Central China | **1.51 (1.11, 2.07)\*\*** | **1.46 (1.10, 1.94)\*** | 1.12 (0.83, 1.51) | **1.41 (1.04, 1.92)\*** |
| Southwest China | **1.78 (1.30, 2.45)\*\*\*** | 1.25 (0.93, 1.69) | 1.06 (0.76, 1.48) | 1.38 (0.98, 1.95) |
| Northwest China | **1.95 (1.46, 2.60)\*\*\*** | **1.54 (1.17, 2.02)\*\*** | **1.70 (1.28, 2.25)\*\*\*** | **1.87 (1.39, 2.53)\*\*\*** |

CI, confidence interval; OR, odds ratio; Ref, reference.
\*\*\**p* < 0.001
\*\**p* < 0.01
\**p* < 0.05.

Greater than 60 years old individuals had a higher risk of having depressive symptoms compared to 18-30 years old in spring (OR _adjusted_ = 1.71, 95%CI = 1.23, 2.36, *p* = 0.001) and winter (OR _adjusted_ = 1.77, 95%CI = 1.27, 2.45, *p* = 0.001) and compared to 46-60 years old in spring (OR _adjusted_ = 1.48, 95%CI = 1.11, 1.98, *p* = 0.007). For individuals born in spring, elementary education was a higher risk factor of having depressive symptoms compared to higher education (OR _adjusted_ = 1.47, 95%CI = 1.05, 2.05, *p* = 0.024). Yet, individuals born in summer with secondary (vs higher) education had a lower risk of having depressive symptoms (OR _adjusted_ = 0.69, 95%CI = 0.50, 0.96, *p* = 0.027).

In the variable of geographic regions, South China was set as the reference. Living in other regions excluding North China, people born in spring had a higher risk in having depressive symptoms. Central and Northwest China showed a higher risk in individuals born in summer. Northeast and Northwest China showed a higher risk in individuals born in autumn. Central and Northwest China showed a higher risk in individuals born in winter. Their ORs and 95%CIs were presented in **Table 2**.

### 3.4 Secondary analyses

While individuals born in June and December reached the highest proportion in having depressive symptoms, individuals born in January and September had lower proportion. However, the rates of having depressive symptoms in each birth month showed no significance difference (given in **Fig. S1**).

We further analysed the influence of seasonality on having depressive symptoms. In line with the primary outcome in **Table 1**, no significant association was observed between season of birth and depressive symptoms (χ^2^ = 3.75, *p* = 0.290, **Table S1**). We also conducted a multivariate binary logistic regression with resampling by the secondary definition of season of birth.

As displayed in **Table S2**, the results of satisfaction of family income remained robust, while the results of gender changed largely. The linear *p* value of gender attributing to having depressive symptoms was not significant in individuals born in summer nor winter. Compared with males, females were still more likely to have depressive symptoms when born in spring (OR _adjusted_ = 1.39, 95%CI = 1.03, 1.87, *p* = 0.031), summer (OR _adjusted_ = 1.34, 95%CI = 1.00, 1.78, *p* = 0.048), and autumn (OR _adjusted_ = 1.41, 95%CI = 1.07, 1.86, *p* = 0.014) but not in winter (OR _adjusted_ = 1.16, 95%CI = 0.83, 1.61, *p* = 0.385).

The significant association between age and depressive symptoms across the four seasons of birth did not change considerably. The results regarding education changed largely. In particular, only the result of individuals born in winter was significant, i.e., elementary education (OR _adjusted_ = 0.59, 95%CI = 0.35, 0.99, *p* = 0.047) and secondary education (OR _adjusted_ = 0.76, 95%CI = 0.53, 1.09, *p* = 0.010) showed a lower risk of having depressive symptoms compared to higher education. Regarding geographical regions, individuals living in Northeast and Southwest China remained a higher risk of depressive symptoms in season of birth over spring compared with individuals living in South China. People living in Central China remained a higher risk in season of birth over summer. As well, people living in Northwest China remained a higher risk in season of birth over autumn and winter (**Table S2**).

## 4. Discussion

China spans over an enormous geographical area, making the country extremely diverse in terms of both population and climate (25). For example, there is a great seasonal and regional difference with the former affecting early life by interfering with mood and hormonal level of pregnant women (20, 33-35). In the same vein, the food supply and dietary pattern also change with the seasons (36-38). As a result, seasons may influence the development of individuals in early life. Particularly, many evidences have highlighted the effects of birth seasonality on depressive symptoms. On possible route by which birth seasonality influence depression is through demographic or biological factors of adulthood depressive symptoms.

Hence, the present study explored the influence of season of birth on the susceptibility of demographic characteristics to adulthood depressive symptoms. Consistent with previous literature (39, 40), we found that females, elders, low education, low satisfaction of family income, and living in the northern geographic region of China were the significant susceptible factors of having depressive symptoms. While the season of birth had no significant association with depressive symptoms, further analysis suggested that the sensitivity of demographic variables to depressive symptoms was different according to people’s birth seasonality. In particular, gender and satisfaction of family income were stable in having depressive symptoms but others were not. However, season of birth did interfere the sensitivity of age, education and region in having depressive symptoms.

Firstly, we found that season of birth was not significant related to adulthood depressive symptoms. Specifically, the analysis indicated that season of birth had no direct effects on depressive symptoms as an external factor during early life. We therefore compared the sensitivity of demographic factors on depressive symptoms in four seasons of birth groups.

Secondly, we found that gender and satisfaction of income level were stable factors in having depressive symptoms. Females showed higher risk in having depressive symptoms than male individuals, which is in line with previous studies (41, 42). In general belief, female individuals are more likely to be depressed than males (43) and often experience a so-called “sandwiched life” between household and work (44). Besides, women tend to be more emotional sensitive when facing such problems (45). Hence, females endure much more pressure in daily life. Besides, satisfaction of family income level was also strongly associated with adulthood depressive symptoms across birth seasonality. Many surveys have similarly reported that lower-income level is an important risk factor in adulthood depressive symptoms, particularly in the Asian countries (46). Due to economic pressure from childcare and daily expenses among other things, many adults become anxious about their financial status. As a subject financial factor, satisfaction of family income cannot only reflect one’s financial status but also self-evaluated society status (47, 48). Our results suggested that satisfaction of family income status plays a key role in having depressive symptoms.

However, the sensitivity of age on depressive symptoms changed across different season of birth groups. Only for people born in spring and winter, age was significantly related to depression. Mainly in 46-60 or >60 years old layers, the difference of age on depressive symptoms across four seasons of birth groups was significant. Those people were mainly born in an era of material deprivation. At the times, their standard of living was relatively low. Besides, the refrigerator has not yet entered the ordinary family household, which implied that food could not be well preserved. The seasons might therefore affect the food supply in people’s daily life. The shortage of food in winter and early spring might greatly influence maternal nutrition. Besides, seasonal infectious diseases in spring may also affect the health status of individuals. These reasons might explain the increased risk of having depressive symptoms among people born in spring and winter.

As for education, we found that higher educated individuals showed lower risk in having depressive symptoms. The results were similar with previous study from China (39, 49). As a categorical variable, geography had different associations with depressive symptoms across birth seasonality. Compared with southwest China, people resident in the northeast and northwest China were more likely to be depressed. The regional climate is commonly dry and cold in north area of China. Besides, their average income of individuals is comparatively low than in other areas of China (39). It has been demonstrated that lower income, low humidity and cold-weather are possible risk factors of depression (50, 51). These might cause the prevalence of depressive symptoms of individuals resident in the north area of China.

In the secondary analysis, the results suggested that the significance of all the demographic characteristics in depressive symptoms changed with birth seasonality. We found that the sensitivity of gender on depression changed greatly in the secondary analysis. The results suggested the result of gender was not robust. Above all, the present study evidenced the influence of season of birth on the demographic factors on adulthood depressive symptoms.

Previous reports mainly focused on the relationship between seasons of birth on physical diseases or severe mental disorders like schizophrenia (25, 52, 53). In addition, most studies investigating the effects of season of birth on depression were from a patient population with a relatively modest number of samples (21, 22, 54). More importantly, the trends of adulthood depressive symptoms in different seasons of birth have seldom be reported from China. Compared with the previous studies, the sampling method was more scientific and systemic in our work. In particular, we investigated individuals from 29 provinces in China with a valid number of 16,185. The population in the present research was therefore large and representative. The relation between season of birth and depressive symptoms were thoroughly discussed. We put forward a hypothesis that season of birth might interfere the effects of demographic factors on depressive symptoms. Interestingly, we found that season of birth did influence the sensitivity of demographic factors on depressive symptoms (particular in age).

However, there are several drawbacks in our work. Firstly, the present study mainly investigated individuals from the labour-forces in China. Although labourers are the majority in the population, it still limits the extrapolation of our conclusions to the general population. Additionally, the association between season of birth and depressive symptoms was unstable. The conclusions of birth season effects in having depressive symptoms have varied among different countries. Hence, further study may focus on comparing the season of birth effects across different populations. Many scholars have put forwarded possible explanations about the effects of birth season on depression, such as maternal nutrition, seasonal infectious diseases, genetics, and hormone levels (22, 23, 55-57). However, the real mechanisms of the birth seasonality effects on depressive symptoms remain unclear. Stronger shreds of evidence may need animal experiments or genetic researches about the effects of birth seasonality on depression. Lastly, the investigation of the present work was cross-sectional, meaning that completely causal explanations are questionable. In order to testify the influence of birth seasonality on demographic factors of depression, future studies should optimally include a more representative sample of the general population in a longitudinal design.

## 5. Conclusion

To summarize, we found that season of birth had no direct effects on depressive symptoms. However, season of birth might influence the demographic risk of adulthood depressive symptoms, especially age. As many reports have evidenced birth seasonality effects on human development and biological indicators, we believe that our phenotypical findings may further help identify the potential early life biomarkers and mechanisms in adulthood depression. Conclusively, this work provides new insights on the demographic risk of depressive symptoms and its importance for future prevention strategies.

## Data Availability

Data analysed in this manuscript were obtained from the CLDS project. The raw data may not be shared by third parties due to ethics requirements, but can be obtained via contacting Center for Social Survey, Sun Yat-sen University in Guangzhou, China. (please refer to http://css.sysu.edu.cn for more information about the CLDS data)

## Acknowledgments

Thank all the volunteered participants and staff members for taking part in the CLDS project. Thank all co-authors for their work in this study.

## Declarations

The authors declare that they have no conflicts of interest in this work.

## Ethics statement

This study used public data CLDS 2016. Reanalysis and this project have been permitted through the ethics committee of the host university.

## Data availability

Data analysed in this manuscript were obtained from the CLDS project. The raw data may not be shared by third parties due to ethics requirements, but can be obtained via contacting Center for Social Survey, Sun Yat-sen University in Guangzhou, China.

## Funding

This research did not receive any specific grant from funding agencies in the public, commercial, or not-for-profit sectors.

## Contributors

Hao. Z: Conceptualization, formal analysis, visualization, and writing - original draft, reviewing, and editing;

Danni. P: Writing - original draft, reviewing, and editing;

Juan. C: Writing - reviewing and editing;

Dong. S: Data curation and resources;

Bin. W: Conceptualization, data curation, resources, formal analysis, methodology, supervision, visualization, writing - original draft, reviewing, and editing.

---

**Figure S1.**
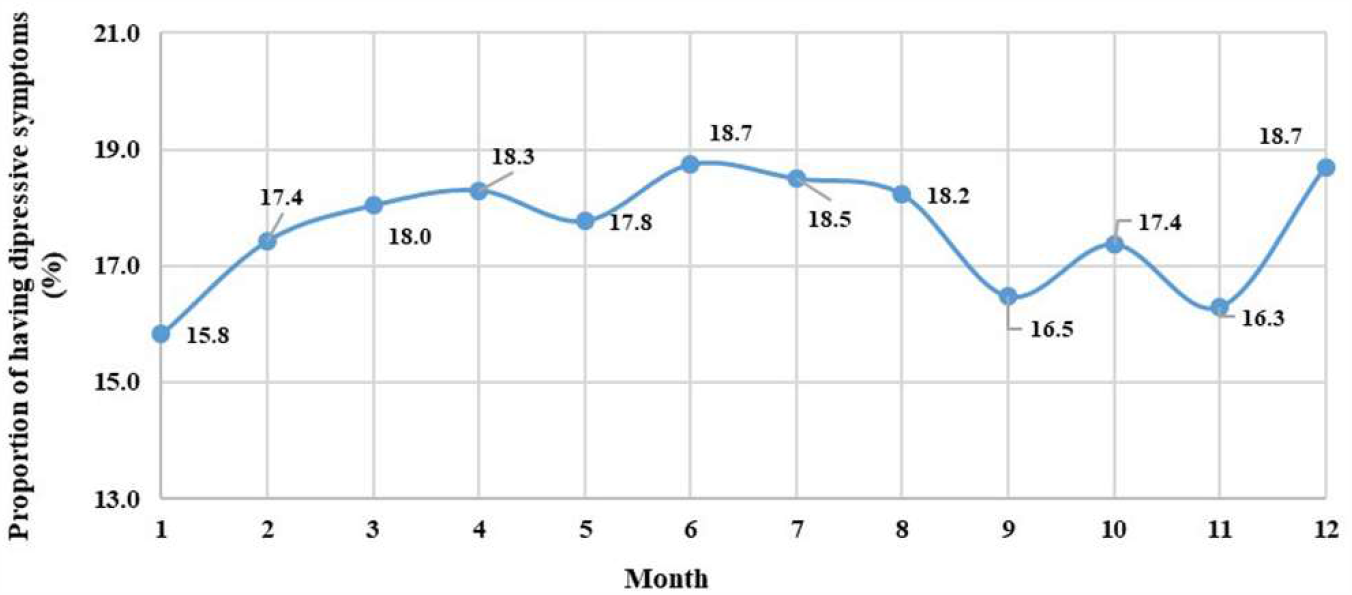
The proportion of having depressive symptoms in each birth month.

**Table S1.** Association of season of birth with depressive symptoms.

| Variables | Non-depressive symptoms |  | Depressive symptoms |  | Chi-square | *p* |
| --- | --- | --- | --- | --- | --- | --- |
|  | N | Proportion | N | Proportion |  |  |
| Season of birth |  |  |  |  | 3.75 | 0.290 |
| Spring | 1,045 | 81.7% | 234 | 18.3% |  |  |
| Summer | 1,107 | 81.5% | 251 | 18.5% |  |  |
| Autumn | 1,327 | 82.6% | 279 | 17.4% |  |  |
| Winter | 984 | 84.3% | 185 | 15.8% |  |  |

**Table S2.** The secondary analysis to predict having depressive symptoms across four seasons of birth.

| Variables | Spring | Summer | Autumn | Winter |
| --- | --- | --- | --- | --- |
|  | OR<sub>adjusted</sub> (95%CI) | OR<sub>adjusted</sub> (95%CI) | OR<sub>adjusted</sub> (95%CI) | OR<sub>adjusted</sub> (95%CI) |
| **Gender**<br>(ref: male) | *p* for trend = **0.028** | *p* for trend = 0.051 | *p* for trend = **0.012** | *p* for trend = 0.382 |
| Female | **1.39 (1.03, 1.87)\*** | **1.34 (1.00, 1.78)\*** | **1.41 (1.07, 1.86)\*** | 1.16 (0.83, 1.61) |
| **Age**<br>(ref: 18-30 y) | *p* for trend = **0.021** | *p* for trend = 0.583 | *p* for trend = 0.116 | *p* for trend = **0.017** |
| 31-45 y | 0.94 (0.55, 1.62) | 0.90 (0.55, 1.49) | 1.14 (0.70, 1.85) | 1.27 (0.74, 2.19) |
| 46-60 y | 1.35 (0.81, 2.26) | 0.86 (0.54, 1.40) | 1.22 (0.76, 1.96) | 1.53 (0.89, 2.62) |
| > 60 y | **1.87 (1.04, 3.34)\*** | 1.12 (0.65, 1.94) | **1.71 (1.11, 2.93)\*** | **2.60 (1.41, 4.80)\*\*** |
| **Education**<br>(ref: Higher) | *p* for trend = 0.750 | *p* for trend = 0.385 | *p* for trend = 0.750 | *p* for trend = **0.040** |
| Elementary | 1.26 (0.71, 2.24) | 1.17 (0.71, 1.94) | 1.01 (0.62, 1.63) | **0.59 (0.35, 0.99)\*** |
| Secondary | 1.23 (0.65, 2.33) | 0.87 (0.49, 1.57) | 0.85 (0.49, 1.49) | **0.76 (0.53, 1.09)\*\*** |
| **Satisfaction of family income**<br>(ref: High) | *p* for trend < **0.001** | *p* for trend < **0.001** | *p* for trend < **0.001** | *p* for trend < **0.001** |
| Low | **7.75 (3.79, 15.87)\*\*\*** | **4.12 (2.17, 7.81)\*\*\*** | **7.05 (3.66, 13.57)\*\*\*** | **14.11 (5.41, 36.81)\*\*\*** |
| Middle-low | **2.36 (1.28, 4.37)\*\*** | 1.32 (0.75, 2.33) | **3.32 (1.85, 5.95)\*\*\*** | **7.15 (3.11, 16.43)\*\*\*** |
| Middle | 1.64 (0.92, 2.92) | 1.02 (0.60, 1.73) | **1.98 (1.12, 3.50)\*** | **3.42 (1.50, 7.77)\*\*** |
| Middle-high | 1.26 (0.70, 2.30) | 0.76 (0.43, 1.32) | 1.21 (0.66, 2.20) | 1.95 (0.84, 4.51) |
| **Geographic region**<br>(ref: South China) |  |  |  |  |
| North China | 1.16 (0.62, 2.17) | 1.18 (0.88, 1.67) | 1.45 (0.87, 2.41) | 1.14 (0.57, 2.28) |
| Northeast China | **2.42 (1.35, 4.35)\*\*** | 1.77 (0.93, 3.38) | 1.55 (0.87, 2.79) | 1.00 (0.48, 2.10) |
| East China | 1.29 (0.79, 2.10) | 1.35 (0.87, 2.09) | 1.26 (0.83, 1.92) | 1.23 (0.71, 2.15) |
| Central China | 1.39 (0.80, 2.39) | **1.92 (1.18, 3.13)\*\*** | 1.30 (0.82, 2.08) | 1.17 (0.64, 2.14) |
| Southwest China | **2.39 (1.38, 4.13)\*\*** | 1.42 (0.84, 2.41) | 1.04 (0.59, 1.83) | 1.39 (0.74, 2.63) |
| Northwest China | 1.55 (0.93, 2.61) | 1.28 (0.78, 2.11) | **1.78 (1.13, 2.81)\*** | **2.02 (1.17, 3.47)\*** |

CI, confidence interval; OR, odds ratio; Ref, reference.
\*\*\**p* < 0.001
\*\**p* < 0.01
\**p* < 0.05.

## Notes

### Competing Interest Statement

The authors have declared no competing interest.

### Author Declarations

The epidemiological study using open data needs no clinical registration.

## Reference

1. Hsieh CR, Qin X. Depression hurts, depression costs: The medical spending attributable to depression and depressive symptoms in China. Health Econ. 2018;27(3):525–44.

2. James SL, Abate D, Abate KH, Abay SM, Abbafati C, Abbasi N, et al. Global, regional, and national incidence, prevalence, and years lived with disability for 354 diseases and injuries for 195 countries and territories, 1990–2017: a systematic analysis for the Global Burden of Disease Study 2017. The Lancet. 2018;392(10159):1789–858.

3. Herrman H, Kieling C, McGorry P, Horton R, Sargent J, Patel V. Reducing the global burden of depression: a Lancet-World Psychiatric Association Commission. Lancet. 2019;393(10189):e42–e3.

4. Cao Y, Li W, Shen J, Malison RT, Zhang Y, Luo X. Health-related quality of life and symptom severity in Chinese patients with major depressive disorder. Asia Pac Psychiatry. 2013;5(4):276–83.

5. Zhang Y, Chen Y, Ma L. Depression and cardiovascular disease in elderly: Current understanding. J Clin Neurosci. 2018;47:1–5.

6. You L, Yu Z, Zhang X, Wu M, Lin S, Zhu Y, et al. Association Between Multimorbidity and Depressive Symptom Among Community-Dwelling Elders in Eastern China. Clin Interv Aging. 2019;14:2273–80.

7. Murakami H, Shiraishi T, Umehara T, Omoto S, Takahashi M, Motegi H, et al. Differences in correlations of depression and anhedonia with cardiovascular sympathetic functions during a head-up tilt test in drug-naive Parkinson’s disease patients. Neurol Sci. 2020.

8. Ren X, Yu S, Dong W, Yin P, Xu X, Zhou M. Burden of depression in China, 1990-2017: Findings from the global burden of disease study 2017. J Affect Disord. 2020;268:95–101.

9. Ru J, Ma J, Niu H, Chen Y, Li L, Liu Y, et al. Burden and depression in caregivers of patients with rheumatoid arthritis in China. Int J Rheum Dis. 2019;22(4):608–13.

10. Guo Y, Sun J, Hu S, Nicholas S, Wang J. Hospitalization Costs and Financial Burden on Families with Children with Depression: A Cross-Section Study in Shandong Province, China. Int J Environ Res Public Health. 2019;16(19).

11. Nolen-Hoeksema S, Larson J, Grayson C. Explaining the gender difference in depressive symptoms. J Pers Soc Psychol. 1999;77(5):1061–72.

12. Xu CJ, Wu WJ, Peng-Li D, Xu PL, Sun D, Wan B. Intraday weather conditions can influence self-report of depressive symptoms. J Psychiatr Res. 2020;123:194–200.

13. Alonso Debreczeni F, Bailey PE. A Systematic Review and Meta-Analysis of Subjective Age and the Association with Cognition, Subjective Wellbeing, and Depression. J Gerontol B Psychol Sci Soc Sci. 2020.

14. Pei YL, Cong Z, Wu B. Education, adult children’s education, and depressive symptoms among older adults in rural China. Soc Sci Med. 2020;253.

15. Pan A, Franco OH, Wang Y-f, Yu Z-j, Ye X-w, Lin X. Prevalence and geographic disparity of depressive symptoms among middle-aged and elderly in China. J Affect Disord. 2008;105(1):167–75.

16. Agarwal N, Aiyar A, Bhattacharjee A, Cummins J, Gunadi C, Singhania D, et al. Month of birth and child height in 40 countries. Economics Letters. 2017;157:10–3.

17. Douros K, Fytanidis G, Papadimitriou A. Effect of the month of birth on the height of young adult males. Am J Phys Anthropol. 2019;170(3):447–50.

18. Sohn K. The influence of birth season on height: Evidence from Indonesia. Am J Phys Anthropol. 2015;157(4):659–65.

19. Torrey EF, Rawlings RR, Ennis JM, Merrill DD, Flores DS. Birth seasonality in bipolar disorder, schizophrenia, schizoaffective disorder and stillbirths. Schizophr Res. 1996;21(3):141–9.

20. Tonetti L, Fabbri M, Martoni M, Natale V. Season of birth and mood seasonality in late childhood and adolescence. Psychiatry Res. 2012;195(1-2):66–8.

21. Talarowska M, Blizniewska K, Wargacka K, Galecki P. Birth Month and Course of Recurrent Depressive Disorders in a Polish Population. Med Sci Monit. 2018;24:4169–74.

22. Mino Y, Oshima I, Okagami K. Seasonality of birth in patients with mood disorders in Japan. J Affect Disord. 2000;59(1):41–6.

23. Björkstén KS, Bjerregaard P. Season of birth is different in Inuit suicide victims born into Traditional than into Modern Lifestyle: a register study from Greenland. BMC Psychiatry. 2015;15:147.

24. Joiner TE, Pfaff JJ, Acres JG, Johnson F. Birth month and suicidal and depressive symptoms in Australians born in the Southern vs. the Northern hemisphere. Psychiatry Res. 2002;112(1):89–92.

25. Wang C, Zhang Y. Season of birth and schizophrenia: Evidence from China. Psychiatry Res. 2017;253:189–96.

26. Xu C, Wu W, Peng-Li D, Xu P, Sun D, Wan B. Intraday weather conditions can influence self-report of depressive symptoms. J Psychiatr Res. 2020;123:194–200.

27. Hoebel J, Maske UE, Zeeb H, Lampert T. Social Inequalities and Depressive Symptoms in Adults: The Role of Objective and Subjective Socioeconomic Status. PLoS One. 2017;12(1):e0169764.

28. Zhang J, Wu Z, Fang G, Li J, Han B, Chen Z. Development of the Chinese age norms of CES-D in urban area. Chinese Mental Health Journal. 2010;24(2):139–43.

29. Radloff LS. The CES-D Scale: A Self-Report Depression Scale for Research in the General Population. Applied Psychological Measurement. 1977;1(3):385–401.

30. Radloff LS. The use of the Center for Epidemiologic Studies Depression Scale in adolescents and young adults. Journal of Youth and Adolescence. 1991;20(2):149–66.

31. Hong Y, Li X, Fang X, Zhao R. Depressive symptoms and condom use with clients among female sex workers in China. Sex Health. 2007;4(2):99–104.

32. Gu ZH, Qiu T, Tian FQ, Yang SH, Wu H. Perceived Organizational Support Associated with Depressive Symptoms Among Petroleum Workers in China: A Cross-Sectional Study. Psychol Res Behav Manag. 2020;13:97–104.

33. Ismailova K, Poudel P, Parlesak A, Frederiksen P, Heitmann BL. Vitamin D in early life and later risk of multiple sclerosis-A systematic review, meta-analysis. PLoS One. 2019;14(8):e0221645.

34. Luykx JJ, Bakker SC, Lentjes E, Boks MP, van Geloven N, Eijkemans MJ, et al. Season of sampling and season of birth influence serotonin metabolite levels in human cerebrospinal fluid. PLoS One. 2012;7(2):e30497.

35. Tonetti L, Milfont TL, Tilyard BA, Natale V. Month of birth and mood seasonality: a comparison between countries in the northern and southern hemispheres. Psychiatry Clin Neurosci. 2013;67(3):133–8.

36. Stelmach-Mardas M, Kleiser C, Uzhova I, Peñalvo JL, La Torre G, Palys W, et al. Seasonality of food groups and total energy intake: a systematic review and meta-analysis. Eur J Clin Nutr. 2016;70(6):700–8.

37. Zhang QF, Lyu CP, Zhou JC, Li XJ, Zheng Y, Xiong C, et al. [Nutrient and metabolic responses of the leaves of Cunninghamia lanceolata seedlings to warming and reduced precipitation in different season]. Ying Yong Sheng Tai Xue Bao. 2019;30(2):420–8.

38. Zhu Z, Wu C, Luo B, Zang J, Wang Z, Guo C, et al. The Dietary Intake and Its Features across Four Seasons in the Metropolis of China. J Nutr Sci Vitaminol (Tokyo). 2019;65(1):52–9.

39. Qin X, Wang S, Hsieh C-R. The prevalence of depression and depressive symptoms among adults in China: Estimation based on a National Household Survey. China Economic Review. 2018;51:271–82.

40. Wu H, Li H, Ding Y, Jiang J, Guo P, Wang C, et al. Is triglyceride associated with adult depressive symptoms? A big sample cross-sectional study from the rural areas of central China. J Affect Disord. 2020;273:8–15.

41. Salk RH, Hyde JS, Abramson LY. Gender differences in depression in representative national samples: Meta-analyses of diagnoses and symptoms. Psychol Bull. 2017;143(8):783–822.

42. Culbertson FM. Depression and gender. An international review. Am Psychol. 1997;52(1):25–31.

43. Parker G, Brotchie H. Gender differences in depression. Int Rev Psychiatry. 2010;22(5):429–36.

44. Borooah VK. Gender Differences in the Incidence of Depression and Anxiety: Econometric Evidence from the USA. Journal of Happiness Studies. 2010;11(6):663–82.

45. Vafaei A, Ahmed T, Freire Ado N, Zunzunegui MV, Guerra RO. Depression, Sex and Gender Roles in Older Adult Populations: The International Mobility in Aging Study (IMIAS). PLoS One. 2016;11(1):e0146867.

46. Gero K, Kondo K, Kondo N, Shirai K, Kawachi I. Associations of relative deprivation and income rank with depressive symptoms among older adults in Japan. Soc Sci Med. 2017;189:138–44.

47. Hoebel J, Muters S, Kuntz B, Lange C, Lampert T. Measuring subjective social status in health research with a German version of the MacArthur Scale. Bundesgesundheitsbla. 2015;58(7):749–57.

48. Reitzel LR, Vidrine JI, Li YS, Mullen PD, Velasquez MM, Cinciripini PM, et al. The influence of subjective social status on vulnerability to postpartum smoking among young pregnant women. Am J Public Health. 2007;97(8):1476–82.

49. Lian Q, Zuo X, Mao Y, Luo S, Zhang S, Tu X, et al. Anorexia nervosa, depression and suicidal thoughts among Chinese adolescents: a national school-based cross-sectional study. Environ Health Prev Med. 2017;22(1):30.

50. Davis RE, McGregor GR, Enfield KB. Humidity: A review and primer on atmospheric moisture and human health. Environ Res. 2016;144(Pt A):106–16.

51. Ding N, Berry HL, Bennett CM. The Importance of Humidity in the Relationship between Heat and Population Mental Health: Evidence from Australia. PLoS One. 2016;11(10):e0164190.

52. Troisi A, Pasini A, Spalletta G. Season of birth, gender and negative symptoms in schizophrenia. Eur Psychiatry. 2001;16(6):342–8.

53. Soreca I, Cheng Y, Frank E, Fagiolini A, Kupfer DJ. Season of birth is associated with adult body mass index in patients with bipolar disorder. Chronobiol Int. 2013;30(4):577–82.

54. Park SC, Sakong JK, Koo BH, Kim JM, Jun TY, Lee MS, et al. Potential Relationship between Season of Birth and Clinical Characteristics in Major Depressive Disorder in Koreans: Results from the CRESCEND Study. Yonsei Med J. 2016;57(3):784–9.

55. Schnittker J. Season of birth and depression in adulthood: Revisiting historical forerunner evidence for in-utero effects. SSM - population health. 2018;4:307–16.

56. Chotai J, Serretti A, Lattuada E, Lorenzi C, Lilli R. Gene-environment interaction in psychiatric disorders as indicated by season of birth variations in tryptophan hydroxylase (TPH), serotonin transporter (5-HTTLPR) and dopamine receptor (DRD4) gene polymorphisms. Psychiatry Res. 2003;119(1-2):99–111.

57. Levitan RD, Masellis M, Lam RW, Kaplan AS, Davis C, Tharmalingam S, et al. A birth-season/DRD4 gene interaction predicts weight gain and obesity in women with seasonal affective disorder: A seasonal thrifty phenotype hypothesis. Neuropsychopharmacology. 2006;31(11):2498–503.

